# Wrist Posture Unpredictably Affects the Perception of Transcutaneous Electrical Nerve Stimulation

**DOI:** 10.1101/2023.11.07.23298213

**Authors:** Neha Thomas, Luke Osborn, Courtney Moran, Matthew Fifer, Breanne Christie

## Abstract

**Objective:** Targeted transcutaneous electrical nerve stimulation (TENS) is a non-invasive neural stimulation technique that involves activating sensory nerve fibers to elicit tactile sensations in a distal, or referred, location. Though TENS is a promising approach for delivering haptic feedback for those with somatosensory deficits, it was not known how the perception of TENS might be influenced by changing wrist position during sensorimotor tasks.

**Approach:** We worked with 12 able-bodied individuals and delivered TENS by placing electrodes on the wrist, thus targeting the ulnar and median nerves, and eliciting tactile sensations in the hand. We recorded perceptual data across three wrist postures: neutral, 45° extension, and 45° flexion. For each posture, the participants drew where they perceived the elicited percepts on a map of the hand. They verbally reported the quality of the percepts in their own words. We also varied the pulse amplitude and width of the stimulation to generate a strength-duration curve, from which we extracted the rheobase current and chronaxie time. Linear mixed models were run on the slope and intercept of the linear fit between pulse width and pulse amplitude to investigate effects of gender, posture, and electrode placement.

**Main results:** As wrist posture changed, sensation quality was modulated for half of the participants, and percept location changed for 11/12 participants. There were no major changes in the surface area of the elicited percepts. The rheobase and chronaxie values were influenced by wrist posture, but the direction of these changes varied by participant and therefore the effect was not systematic. The statistical models indicated interactions between posture and electrode placement, as well as an effect of gender.

**Significance:** If using TENS to convey haptic feedback in sensorimotor tasks, in which wrist posture will likely change, it may be important to characterize resulting perceptual changes for individual users.

## Introduction

Haptic feedback is critical for manipulating objects (1) and plays a major role in emotional connection (2). The role of cutaneous touch is especially important when visual feedback is unavailable or unreliable (3). For those living with somatosensory deficits in their hands, it can be difficult to grip objects with an appropriate amount of force that will neither crush the object nor allow it to slip from one’s grasp. With current technological advancements, there are also many scenarios in which we do not have haptic feedback but may benefit from it, such as in telemedicine or in extended reality.

Haptic feedback can be administered via invasive or non-invasive neural interfaces. Electrical stimulation can be delivered at various points along the somatosensory pathway to elicit tactile percepts, which can be used as a source of haptic feedback. Prior studies have delivered electrical stimulation to the somatosensory cortex to elicit artificial touch in people with spinal cord injury (4). Other studies implanted neural interfaces onto the peripheral nerves or spinal cord to elicit tactile feedback in people with amputations (5,6). These approaches can selectively activate distinct populations of neurons, which allows them to activate different regions of the hands. However, invasive neural interfaces are expensive and require surgical intervention, so it is not as appealing for many individuals, particularly when the use-case is not for health purposes.

“Smart gloves” and transcutaneous electrical nerve stimulation (TENS) are two non-invasive methods for delivering haptic feedback. Smart gloves contain vibrating motors, pneumatic chambers, or wire actuators that are placed on a person’s hand (7,8). These devices are intuitive and fairly easy to use, but they can be bulky and constrain natural hand movements and sensations. “Targeted” TENS, however, can elicit tactile feedback in the hand without the physical constraints of a glove. By placing electrodes more proximally, such as on the wrist or biceps, it is possible to target and activate the sensory fibers (i.e., median and ulnar nerves) that innervate the hand.

TENS electrodes placed on the forearm (9), biceps (10), elbow (11), or wrist (12) have successfully elicited tactile sensations in the hands of able-bodied individuals. TENS has been described as feeling like tingling, vibration, pulsing, fluttering, tapping, electricity, or touch (12–14). The perception of TENS obeys a strength-duration curve, in which perception is governed by an inverse relationship between pulse amplitude and pulse width/duration (11,12). As pulse amplitude increases, the pulse width needed to maintain detection of the stimulus decreases. Finally, it is possible to use TENS as a method for conveying feedback about grip. In a prior study, able-bodied individuals received feedback about an object that was being picked up by a prosthetic hand (10). TENS amplitude was scaled with respect to fingertip force. The individuals were able to determine object shape and recognize topological surface features with high levels of accuracy.

It is not yet known if the perception of TENS will be influenced by changing joint angles, which may shift the location of the targeted sensory fibers (15). The stability of TENS perception during dexterous movements will be critical for the utility of TENS in real-time functional tasks. Therefore, the primary goal of this study was to characterize the perception of TENS across different wrist postures. We hypothesized that changes in wrist posture would alter detection thresholds, but would not alter the perceived location, quality, or size of the evoked percept.

## Methods

### Targeted TENS

This study was conducted with 12 able-bodied people (seven male) between 21-41 years old. The study protocol was approved by the Johns Hopkins Medicine Institutional Review Board. Informed consent was obtained prior to participation in research-related activities.

TENS was delivered using a DS8R Biphasic Constant Current Stimulator (Digitimer®, Welwyn Garden City, UK). The electrodes were ∼1 cm in diameter and adhered to the skin using medical tape. The stimulating electrode was placed on the right wrist, and the reference electrode was placed on the metacarpophalangeal joint of the right thumb (Figure 1). The stimulating electrode placement varied by person. At the start of each session, we performed an exploration in which we slowly increased stimulation pulse amplitude while the participant moved the electrode around in a mediolateral direction on the palmar side of their wrist. This exploration was stopped once we identified a location at which stimulation was capable of eliciting a percept in their hand.

**Figure 1:**
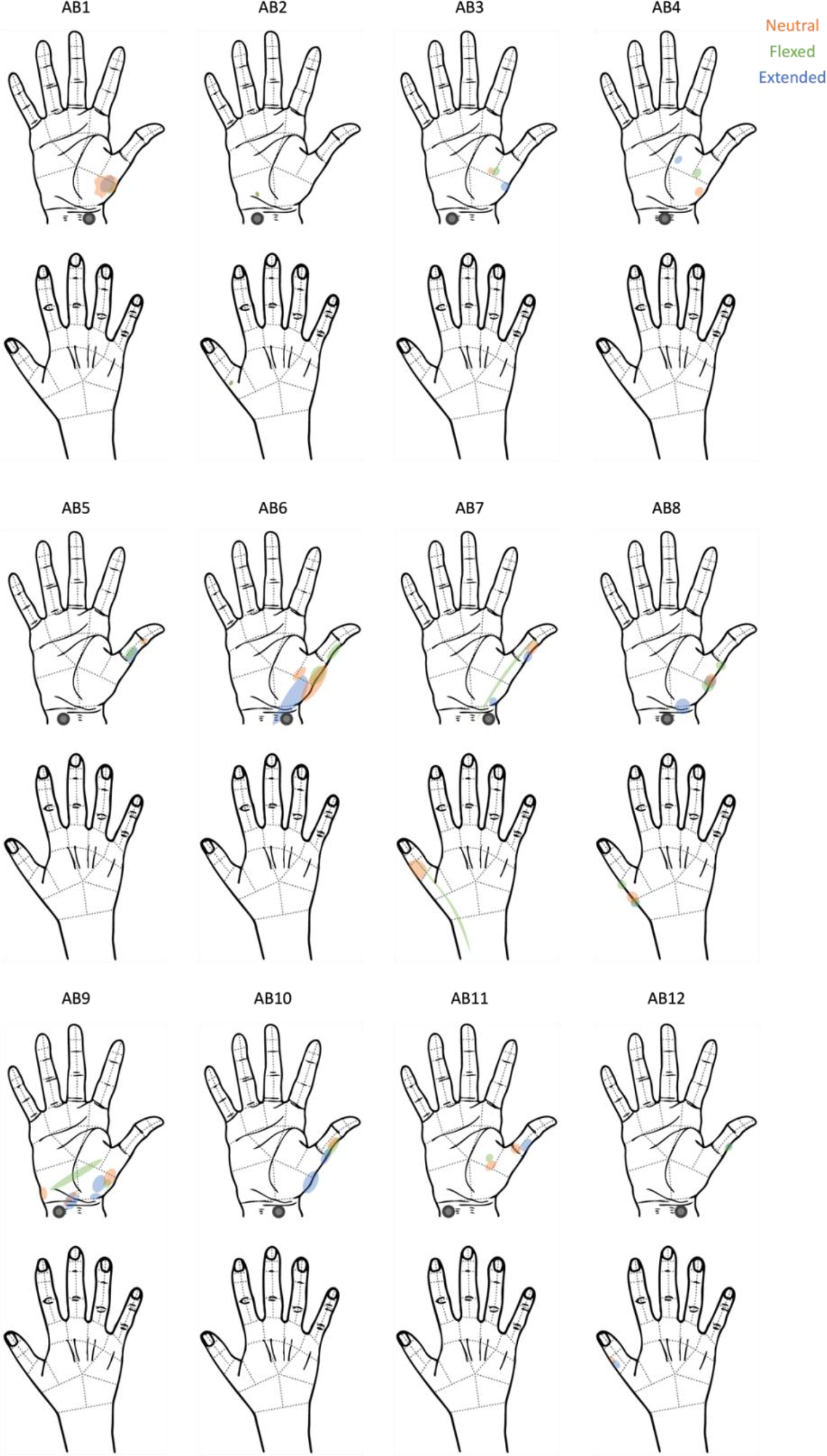
The participants’ reports of where they felt the tactile percepts elicited by TENS. The electrodes were placed on the right wrist, as indicated by the grey circle.

### TENS perceptions in different wrist postures

TENS detection thresholds were estimated with a two-alternative forced-choice (2-AFC) paradigm in which the participant verbally reported which of two 0.5 sec intervals contained a 200 ms stimulus. Auditory cues were used to indicate the start of each interval, with a different pitch assigned to each interval. A 3 down-1 up (3D-1U) adaptive staircase procedure was used to estimate the thresholds (16). We decreased the stimulus intensity until no percept was perceived, at which point a reversal occurred and the intensity increased. Three correct responses in a row at one intensity level led to a decrease in the intensity. The intensity increased after one incorrect answer. When stimulation pulse width (PW) was held constant, the pulse amplitude (PA) was changed with a step size of 0.1 mA. When PA was held constant, PW was changed with a step size of 10 μs or 20 μs. The 10 μs step size was used when the PA was at a higher level, because smaller changes in PW produced greater changes in delivered charge that resulted in more noticeable changes in intensity. The detection threshold was calculated by averaging the stimulus intensity across all reversals (i.e. whenever the direction of the staircase changed). This method estimates the stimulus intensity that can be detected 79.4% of the time (16).

Thresholds were collected while the wrist was held in three different postures: neutral, extended at a 45° angle, and flexed at a 45° angle. For the first six participants, the participants rested their fingertips on a box to maintain this angle. For the remaining six, the participants rested their hand on a 3D-printed 45° ramp. For each posture, we collected three thresholds while holding PW constant at 100, 200, or 500 μs and three thresholds while holding PA constant at 0.75, 1, or 2 mA. For the first three participants, we attempted to hold PA constant at 0.5 mA instead of 0.7 mA; however, we found that the pulse width had to reach closer to 1000 μs to become detectable and it became too difficult to quickly find a threshold. At a suprathreshold stimulation level, the participants were asked to outline where they perceived the TENS percept on a drawing of the hand (**Figure 1**). They were also asked to describe the quality of the percept in their own words. Experiments lasted approximately two hours per person.

The data points collected during threshold testing were fit to a strength-duration curve using Lapicque’s equation (*Equation 1*) (11,12,17–19). Lapicque’s equation was fit to the raw data using a nonlinear least-squares method with bisquare weighting, with starting values of b=0.1 and c=1000. The R-squared goodness of fit values had an average of 0.97. In Lapicque’s equation, b is the rheobase current, c is the chronaxie time, d is the pulse width, and I is the pulse amplitude.

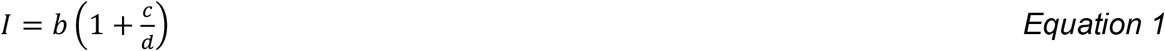

The rheobase corresponds to the lowest pulse amplitude that is able to elicit a detectable percept at an infinitely high pulse width. The chronaxie time is the pulse width that corresponds to twice the rheobase. The resulting curve presents a relationship between the minimum pulse amplitude and pulse width required to elicit a detectable percept. However, because we typically did not test pulse widths higher than 500 μs, the Lapicque equation’s coefficients were not representative of the approximate asymptote. Instead, we approximated that the asymptote was reached near 1000 μs based on a prior study (11), which we deemed to be the rheobase current, and calculated the chronaxie time.

### Statistical Methods

We ran two-tailed paired t-tests between the rheobase amplitudes in the neutral vs. extended and the neutral vs. flexed conditions. We also ran the same comparisons for the chronaxie values. P-values less than 0.05 were considered to be statistically significant.

Lapicque’s equation was transformed into a linear relationship by multiplying by *d*, the pulse width. This linearized form is commonly referred to as Weiss’s equation. Thus, *b* (the rheobase current) was the slope of the line, and *b*c* was the intercept. We ran two linear mixed models on *b* and *b*c. b* was log-transformed when running the mixed model due to non-normal residuals. Covariates of the model included unique combinations of posture, electrode placement, use of the ramp (15 total conditions), and gender. Custom contrasts were applied to the condition variable to make specific comparisons. Raw p-values were multiplied by two to make appropriate corrections for multiple comparisons. The comparisons were as follows:

1. Flexed posture vs. neutral posture
2. Extended posture vs. neutral posture
3. Ulnar electrode vs. radial electrode
4. Ulnar electrode vs. central electrode
5. Flexed posture with central electrode with ramp vs. neutral posture with central electrode
6. Flexed posture with central electrode without ramp vs. neutral posture with central electrode
7. Flexed posture with radial electrode with ramp vs. neutral posture with radial electrode
8. Flexed posture with radial electrode without ramp vs. neutral posture with radial electrode
9. Flexed posture with ulnar electrode with ramp vs. neutral posture with ulnar electrode
10. Extended posture with ulnar electrode with ramp vs. neutral posture with ulnar electrode
11. Extended posture with ulnar electrode without ramp vs. neutral posture with ulnar electrode
12. Extended posture with radial electrode with ramp vs. neutral posture with radial electrode
13. Extended posture with radial electrode without ramp vs. neutral posture with radial electrode
14. Extended posture with central electrode with ramp vs. neutral posture with central electrode

Post-hoc tests included:

1. Flexed posture vs. extended posture
2. Central electrode vs. radial electrode
3. Flexed posture with radial electrode with ramp vs. without ramp
4. Flexed posture with central electrode with ramp vs. without ramp

## Results

The electrodes were placed on the palmar side of the right wrist. The precise locations were chosen via an initial exploration, in which we slowly increased stimulation pulse amplitude while the participant moved the electrode in a mediolateral direction until they felt a tactile percept in the hand. Five individuals had a more ulnar placement (towards the pinky), three had a central placement, and four had a more radial placement (towards the thumb).

Some of the words that individuals used to describe TENS percept qualities were flick, buzz, twitch, pins and needles, burst, tensing, pulse, tingling, muscle spasm, tapping, poking, flick, stroking, pulling, swelling, and vibrating (**Table 1**). The qualities of the reported percepts changed as a result of changing wrist posture for half of the participants. Typically, even though different words were reported, most participants reported that any changes were minor. For example, with participant AB6, tingling was reported in the flexed and neutral wrist postures, and pins and needles were also reported when in the extended posture. Only one participant (AB7) reported a completely different sensation between the neutral and flexed postures.

**Table 1:** The participants’ reports of how tactile percepts elicited by TENS felt.

| Participant ID | Placement of Stimulating Electrode on the Wrist | Quality, Wrist = Extended | Quality, Wrist = Flexed | Quality, Wrist = Neutral |
| --- | --- | --- | --- | --- |
| AB1 | Radial | *Same as neutral* | *Same as neutral* | Twitch, like a fly landed on his hand |
| AB2 | Ulnar | *Same as neutral* | *Same as neutral* | "Ghost twitch" pins and needles, but a single needle |
| AB3 | Ulnar | *Same as neutral* | Burst pattern when stimuli became stronger | Tingling up fingers |
| AB4 | Central | *Same as neutral* | *Same as neutral* | Tensing pulse |
| AB5 | Ulnar | *Same as neutral* | *Same as neutral* | Low grade pins and needles |
| AB6 | Radial | Tingle, pins and needles on top of the tingle | *Same as neutral* | Tingle |
| AB7 | Radial | Less abrupt sensation (shorter in duration) than neutral | Light stroke on hand | Weaker stimulation felt like buzz pulse, stronger felt like someone flicking |
| AB8 | Central | *Same as neutral* | *Same as neutral* | Tapping |
| AB9 | Ulnar | *Same as neutral* | *Same as neutral* | Rippled muscle spasm, felt in sequence (pinky, then thumb) |
| AB10 | Central | Flick, buzz | *Same as neutral* | Flick, got longer when stimuli became stronger |
| AB11 | Ulnar | Buzzy poking | Pulling, swelling | Vibrating, poking, swelling, pulling |
| AB12 | Radial | Pins and needles | Pins and needles | Heartbeat pulse, pins and needles |

We were able to elicit tactile sensations referred to the hand in all participants. The locations of the reported percepts changed as a result of changing wrist posture for all participants except for AB2 (**Figure 1**). The percept size did not vary greatly between wrist postures.

The psychometric data displayed an inverse relationship between pulse amplitude and pulse width (**Figure 2)**, which is typical of a strength-duration curve. The rheobase current was 0.262 ± 0.029 mA (mean ± standard error across participants) in the neutral position, 0.258 ± 0.038 mA in the flexed position, and 0.250 ± 0.028 mA in the extended position (**Figure 3**). The chronaxie time was 465.37 ± 17.86 μs in the neutral position, 475.42 ± 12.77 μs in the flexed position, and 472.04 ± 13.92 μs in the extended position. The rheobase and chronaxie values were not statistically significantly different between the neutral wrist position and the flexed or extended positions (paired t-tests, p>0.05). However, this does not mean that there were no effects of wrist posture on detection thresholds. For five participants (AB-3, 8, 10, 11, 12), the psychometric curves are nearly overlapping for the three wrist postures. However, for the remaining participants, there is some separation between the conditions, but there is not a consistent trend. As an example, for participant AB1, detection thresholds were the highest when the wrist was in the flexed position and highest in the extended position. Conversely, for participant AB2, detection thresholds were the highest when the wrist was in the flexed position. Eight of the 12 participants had lower rheobase amplitudes (i.e., they were more sensitive) in the flexed position compared to the neutral position, though again this was not statistically significantly different. A different subset of eight participants also had lower rheobase amplitudes in the extended position compared to the neutral position.

**Figure 2:**
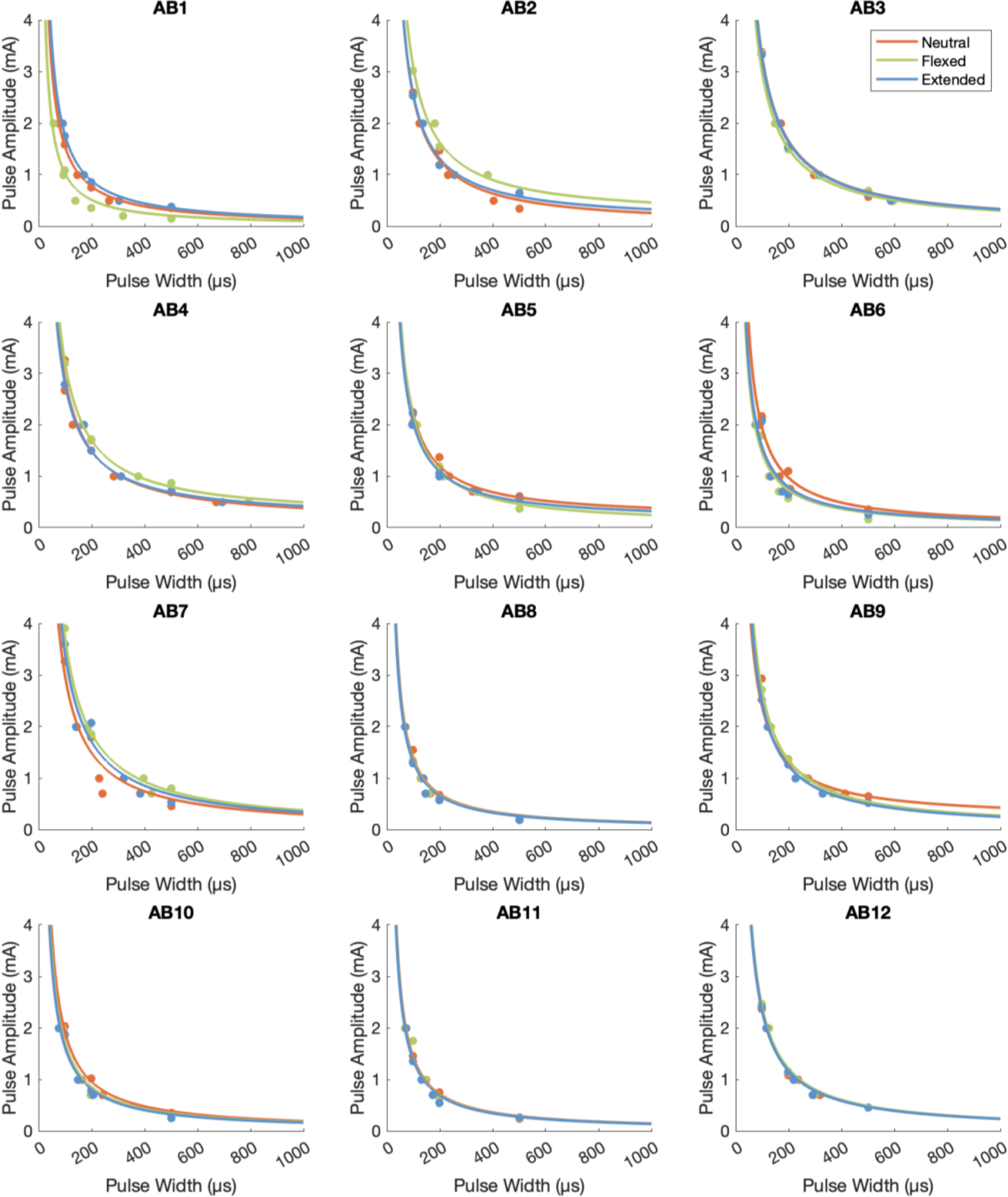
The psychometric data for TENS perception for the twelve able-bodied participants. The raw data points are depicted with dots, and Lapicque’s equation was fit to the data from each posture.

**Figure 3:**
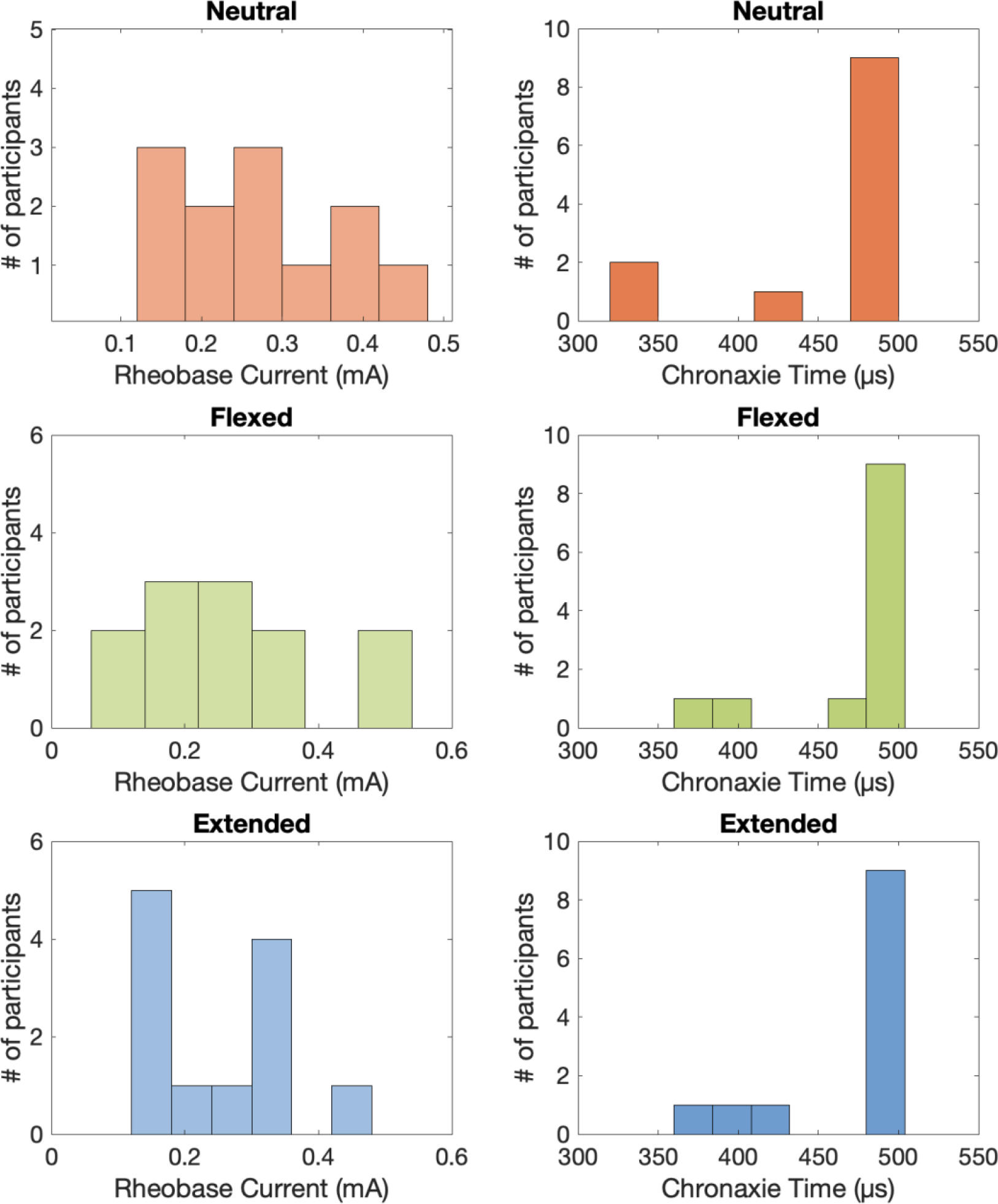
The approximated rheobase currents and chronaxie times for TENS perception.

Results of the linear-mixed models indicate that overall electrode placement on the wrist had an effect on the slope of the linear relationship between pulse width and pulse amplitude, but no effect on the intercept (**Tables 2 and 3)**. Central placement had a steeper slope than radial placement. This indicates that a central electrode placement results in higher thresholds, especially for larger pulse widths, compared to thresholds for an electrode placed more radially.

**Table 2:**
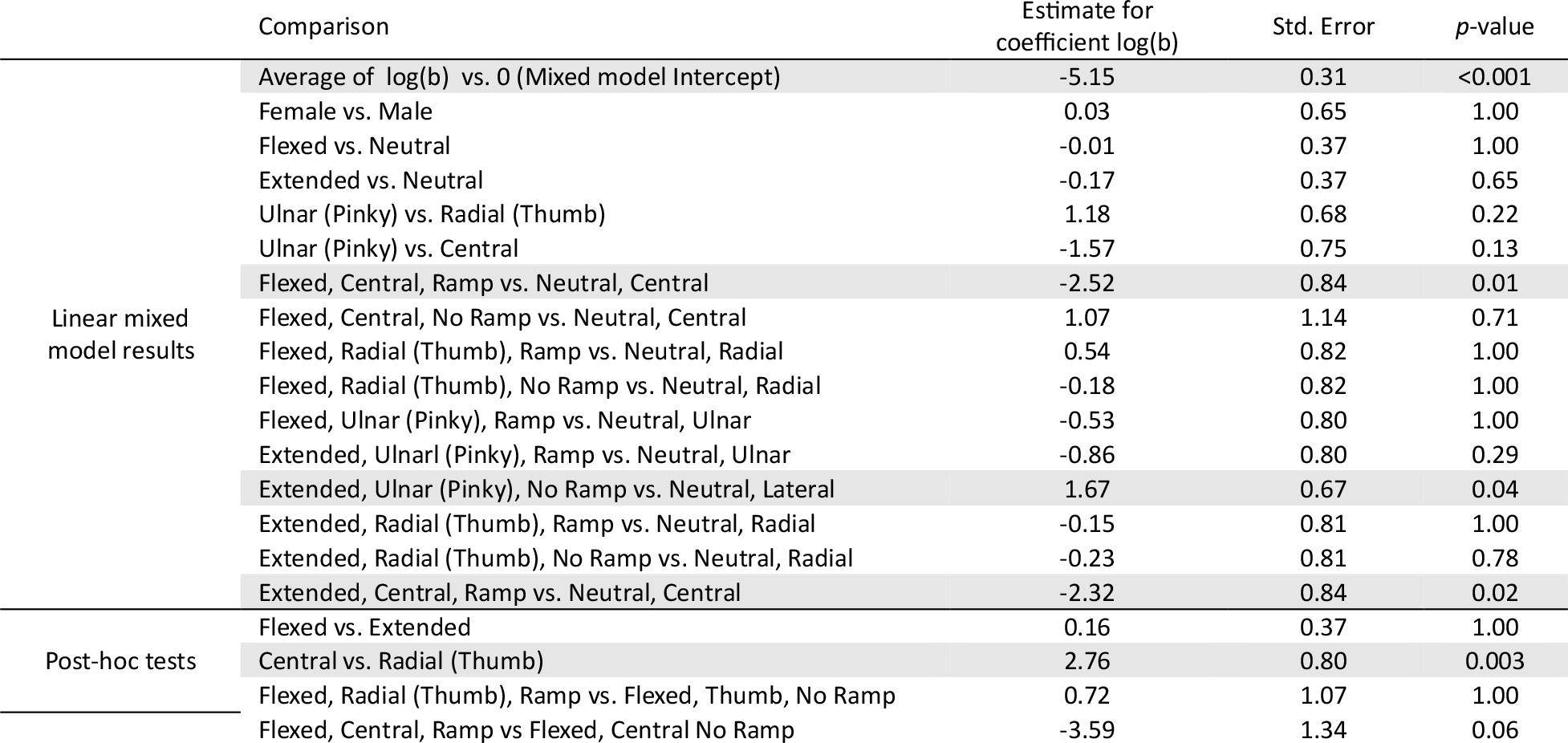
Linear mixed model results for comparisons of the slope of Weiss’s equation (b, the rheobase current), followed by post-hoc tests. Shaded rows indicate p-values less than 0.05.

**Table 3:**
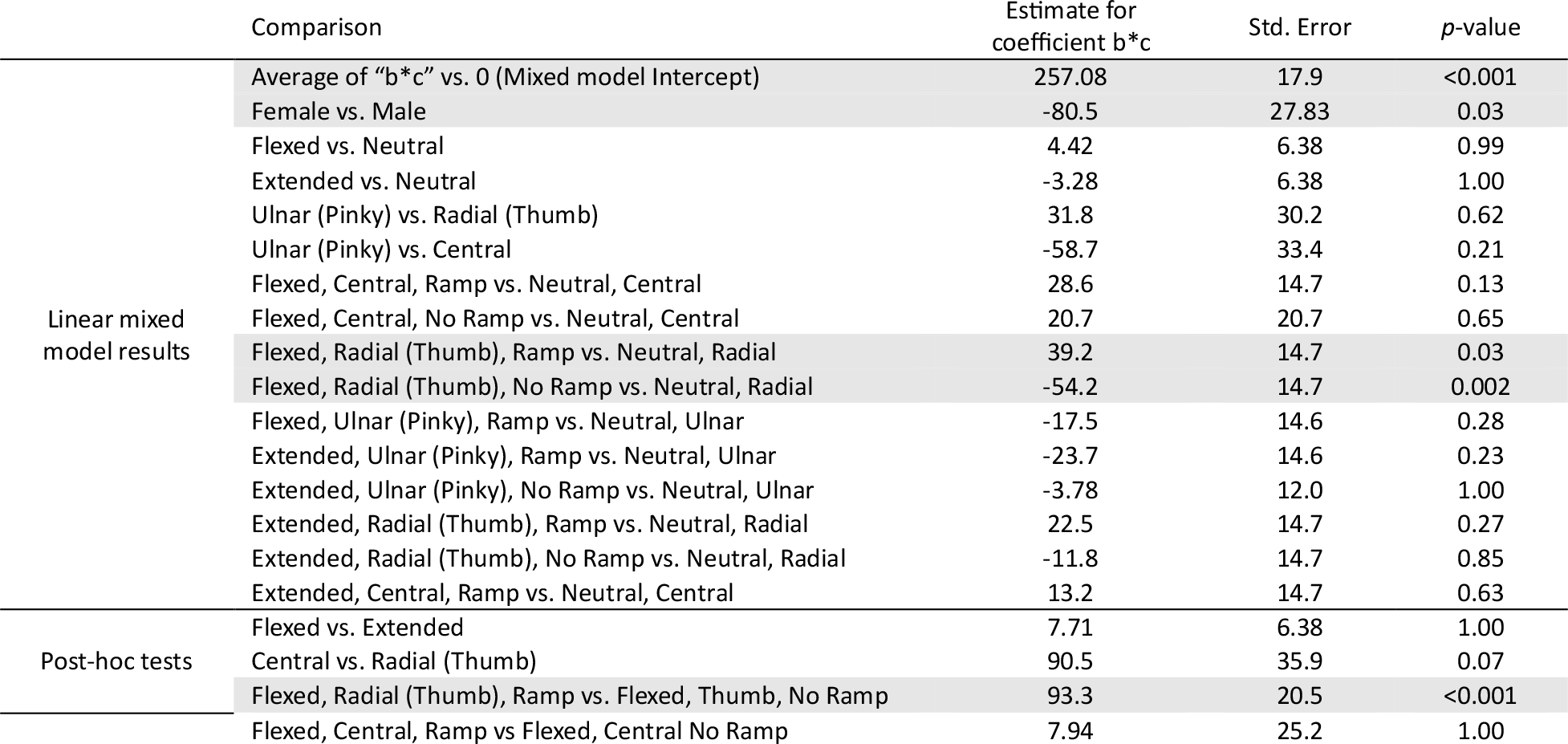
Linear mixed model results for comparisons of the intercept of Weiss’s equation (b*c), followed by post-hoc tests. Shaded rows indicate p-values less than 0.05.

Female participants had similar slopes as male participants, but significantly lower intercepts, indicating overall lower thresholds. Furthermore, particular combinations of posture, electrode placement, and the use of the ramp had significant effects on the slope and intercept. A flexed posture with a central electrode *with* a ramp had a significantly shallower slope than a neutral posture with a central electrode placement. Similarly, an extended posture with a central electrode with a ramp had a significantly shallower slope than a neutral posture with a central electrode. The intercept was significantly higher for flexed posture with radial electrode with a ramp compared to without a ramp. Thus, the ramp in this particular posture and electrode configuration increased thresholds.

## Discussion

The primary objective of this study was to characterize the perception of TENS across wrist postures that are commonly adopted during dexterous movements. Secondary objectives included investigating the effects of stimulating electrode placement on the wrist, utilization of a ramp to enforce flexed or extended postures, and gender. To achieve these objectives, we examined percept size, location, and quality; the chronaxie and rheobase values for each posture; and the intercept and slope of the linear fit between pulse width and pulse amplitude.

Participants reported the tactile percepts as originating distally in their hand rather than in the skin below the electrodes, indicating that our TENS approach was indeed targeted and stimulated underlying sensory fibers that innervated the hand. Consistent with prior studies, TENS was most commonly described as feeling like pins and needles, buzzing, and/or pulsing (12–14). As wrist posture changed, there were minor changes in sensation quality for half of the participants, substantial percept location changes for 11/12 participants, and no major change in percept size for any participants. There was no consistent relationship between the direction of the movement and the hand posture, nor between the direction of the movement and the electrode placement. We believe that changing wrist posture may have shifted the location of the underlying sensory fibers targeted by TENS (15), but that differences in individual anatomy and variations in electrode placement likely prevented there from being a systematic pattern.

There were no statistically significant differences in the rheobase, chronaxie, or coefficients of the fitted curves between the three wrist postures (neutral, extended, and flexed). While rheobase values could vary across postures, the effect was not systematic. Additionally, we found that placement of the stimulating electrode on the wrist significantly influenced only the slope, with a central placement resulting in steeper slopes and therefore higher thresholds. The central placement of the stimulating electrode likely targets the median nerve; the lower sensitivity of the central placement may then be due to neighboring anatomical structures within the carpal tunnel (20).

There were also potential differences in slope when considering the use of a ramp to enforce hand posture. The presence of the ramp increased thresholds for a flexed posture with radial electrode. When no ramp was used, the back of the participants’ fingers rested on the edge of a box, which may have allowed participants the ability to flex more naturally in the ulnar direction, where there is greater range of motion (21), than in the radial direction. When the ramp was used, it enforced flexion to be equal in radial and ulnar directions, which may have impacted sensitivity for a radially placed electrode. Interestingly, both flexion and extension with a central electrode using a ramp resulted in lower thresholds compared to a neutral posture with a central electrode. This may be due to how wrist angle deforms the structures in the wrist, potentially giving better access to the large median nerve than in the neutral posture.

One significant limitation of this study was the curve fitting of the Lapicque’s equation to the raw data. Because we were not able to test pulse widths greater than 500 μs, our values for the rheobase and chronaxie times were only estimates and not the ground truth values based on participant data. Another limitation, which would be interesting to characterize in further experiments, is that we only explored one electrode location per person. It would be interesting in future experiments to systematically vary the electrode placement on the wrist and examine the influence of stimulation location on perception.

Taken together, the results showcase the interacting effects of wrist posture, electrode placement, as well as rigidity of the posture enforced. Future research should collect more data with varying electrode placements and postures, such as placing electrodes on the forearm and studying the influence of elbow flexion. In addition, the perception of TENS during dynamic movements should be assessed. It is possible that while individuals actively perform a task, they may not notice minor changes in percept location, quality, or intensity/detection.

## Conclusion

In conclusion, when placing TENS electrodes on the wrist, wrist posture impacted the perception of tactile percepts that were elicited in the hand, but there was not a consistent trend across participants. If using TENS as tactile feedback in sensorimotor tasks, in which wrist posture will likely change, it may be important to characterize perceptual changes for individual users. Our results suggest that placing the stimulating electrode more centrally on the wrist, rather than towards the ulnar or radial side, may be a more robust feedback method across multiple hand postures, regardless of whether they are more actively or passively held. Overall, though we did observe an impact of wrist posture, TENS remains a promising technique for delivering haptic feedback.

## Data Availability

All data produced in the present study are available upon reasonable request to the authors.

## Acknowledgments

The authors would like to thank the research participants for their time, patience, and dedication. This research was supported by internal funding from the Johns Hopkins University Applied Physics Laboratory.

